# Cognitive mechanisms of confabulations in Alzheimer’s Disease

**DOI:** 10.64898/2026.01.30.26345197

**Authors:** Geoffroy Gagliardi, Valentina La Corte, Marion Houot, Bruno Dubois, Gianfranco Dalla Barba

## Abstract

**Background:** Alzheimer’s disease (AD) patients are characterized by an early decline of episodic memory due to hippocampal damage. Nonetheless, besides the classical negative symptoms related to episodic memory deficits, i.e. failure to retrieve information, it has been shown that AD patients can also suffer from positives symptoms, i.e. confabulations. Some theoretical accounts have been proposed to explain the cognitive mechanisms underlying confabulation. Yet, even if most of these models have lead to some research trying to validate cognitive deficits in some cognitive domains, in particular executive functions, to our knowledge, none has yet tried to determine the specific cognitive profile of confabulatory patients. In the present study the main aim is to characterize the specific cognitive profile of confabulatory patients. Thus, given that AD patients’ cognitive profile is well known and documented, we compare mild to moderate AD patients with and without confabulations.

**Methods:** 37 healthy control (HC) and 35 individuals with mild to moderate AD were recruited at the Pitié Salpêtrière University Hospital. All participants were evaluated on Dalla Barba’s Confabulation Battery to determine their tendency to produce provoked confabulations. Thus, among AD patients, we distinguish between those who produced confabulations in episodic memory questions, and those who did not. Accordingly 27 AD patients were considered free of confabulations (ADC-), and 8 as confabulators (ADC+) (none HC met the criteria). All participants were assessed on a comprehensive neuropsychological battery.

**Results:** Statistical analyses showed a significant difference between HC participants and the two groups of AD patients, in almost all cognitive domains assessed. However, when comparing the two AD groups, they did not show distinct profiles. Moreover, regarding the type of confabulations, ADC+ produced significantly more confabulations to the Episodic questions (both concerning past and future).

**Conclusions:** By not demonstrating cognitive differences between patients with and without confabulations, our results cast doubts on some confabulation models, which assume a unique and sufficient cognitive (e.g. executive) deficit underlying the onset of confabulations.

**Highlights:**

- Alzheimer’s disease patients with or without confabulations do not have otherwise distinct cognitive profiles.
- The emergence of a confabulatory syndrome does not seem to be the result of a necessary and sufficient executive deficit
- Alzheimer’s disease patients mainly produce episodic memory confabulations, which involve both the past and the future dimension.

## 1. Introduction

Episodic Memory (EM) is defined as the ability to store and retrieve personally experienced events associated with a spatial and temporal context (Eustache and Desgranges, 2008; Tulving, 1972, 1985; Wheeler et al., 1997). Authors have determined three processes in EM associated with different neural substrates: encoding, consolidation/storage, and retrieval (Tromp et al., 2015).

Some authors consider that encoding and retrieval processes strongly depend on executive functioning components. Regarding anatomical data, authors thus demonstrated a significant implication of prefrontal cortex (PFC) in these operations (Habib et al., 2003). The episodic retrieval is described as a conscious mental time travel (Eustache and Desgranges, 2008; Mahr and Csibra, 2017; Tulving, 1972), which also allow us to project ourselves in an episodic future (e.g. to plan and anticipate) (Wheeler et al., 1997). The storage/consolidation of the memory trace would involve the medial-temporal lobe (MLT) regions, and especially the hippocampal structures (Dickerson and Eichenbaum, 2010; Kesner, 2013; Langston et al., 2010).

EM can be affected by omissions (Schacter, 2001) or “negative” symptoms (Attali et al., 2009; Dalla Barba and Wong, 1995; Dalla Barba et al., 1995), and commissions (Schacter, 2001) or “positive” symptoms of EM (Attali et al., 2009; Dalla Barba and Wong, 1995; Dalla Barba et al., 1995). The first represents the failure to retrieve desired information, the second concerns memory distortions that can occur during retrieval or recognition phases. Among these positive signs, confabulations are defined as a memory distortion leading to statements or actions incongruent with one’s past, present and future (Dalla Barba, 1993, 2002; Kopelman, 2010; Metcalf et al., 2007). Erroneous memories can either be false or real, but chronologically misplaced events (Dalla Barba and La Corte, 2013). Usually encountered following lesions of the dorsomedial nucleus of the thalamus in Korsakoff’s syndrome, confabulations can also be found after lesions in more than 20 different brain areas (Dalla Barba and Boisse, 2010), following many etiologies (Dalla Barba and Decaix, 2009; Dalla Barba and La Corte, 2015), or even in normal subjects (Burgess and Shallice, 1996; Dalla Barba, 2002; Kopelman, 1987). There is no specific and unique brain lesion site associated to confabulation. However, some authors have observed that confabulation is frequent after injuries in ventromedial and orbitofrontal cortices (Kopelman, 2010). Yet, based on a theoretical and anatomical review, other authors pointed out that, for confabulation to occur, the hippocampus needs to be at least partially spared (Dalla Barba and La Corte, 2013).

Several models have been proposed to account for confabulations. They can be regrouped in at least three categories (Glowinski et al., 2008; Metcalf et al., 2007): those that suppose an executive impairment, those that trace back confabulation to a temporal ordering deficit, and multi-factorial models that consider confabulation the result of multiple cognitive deficits. Regarding the executive models of confabulations, authors postulate two possible processes to be involved in confabulation: source monitoring (Johnson and Raye, 1998; Johnson et al., 1993) and strategic retrieval (Moscovitch and Melo, 1997). The source monitoring approach postulates that confabulatory patients could experience some difficulties to discriminate between reality (true memories) and self-originated episodes (e.g. dreams and imagination), and to ‘source’ in a trustworthy way the temporo-spatial context of memories (Johnson and Raye, 1998; Johnson et al., 1993). The strategic-retrieval approach assumes, at least, three possible failures: error in the search processes, in the association between external cues and memory representations, or in the monitoring leading to faulty acceptance of erroneously activated memory traces (Gilboa, 2010).

The ‘temporal’ approach considers the confusion about chronology to be primarily involved in confabulation (Dalla Barba, 2002; Schnider et al., 1996). In particular, the “Memory, Consciousness and Temporality Theory” (MCTT) (Dalla Barba, 2002) considers confabulation as a distortion of one’s general ability to phenomenologically experience the remembering of the personal past, to be oriented in the present and to project in a personal future. Confabulatory patients would then be unable to make a distinction between a specific and contextualized isolated event on the one hand, and more stable patterns of modifications of the nervous system, i.e. repeated and over-learned information, on the other hand (Dalla Barba and Boisse, 2010; Dalla Barba and Kopelman, 2017; La Corte et al., 2011a, 2011b).

Finally, the multi-factorial approach proposes imprecise explanation regarding the cognitive bases of confabulations (Johnson et al., 1997; Shapiro et al., 1981).

Even if most of these models has lead to research trying to validate cognitive associations with isolate tests, there is still an open debate to determine the cognitive origin of confabulations (e.g. whether this symptom could be explained by an executive, EM, or global cognitive deficit). Moreover, as far as we know, few have yet tried to determine the cognitive profile of confabulatory patients (e.g. Dalla Barba et al., 1999; Nedjam et al., 2004). Our work proposes to determine the global cognitive profile of confabulatory patients, opposing them to patients free from confabulations. One difficulty is the variety of confabulation’s etiologies (Dalla Barba and Boisse, 2010; Dalla Barba and Decaix, 2009; Dalla Barba and La Corte, 2015). This could be solved selecting confabulating patients suffering from the same pathology. We thus selected participants with mild to moderate Alzheimer’s Disease (AD). AD is a neurodegenerative pathology characterized by a progressive atrophy, classically beginning in the hippocampus (Apostolova et al., 2010; Frisoni et al., 2008; Jack et al., 2016; Khan et al., 2015). The cognitive, and EM, profile of AD patients is well-known. First, patients experience EM deficits, followed by executive, attentional, and possibly a general cognitive decline (Dubois et al., 2007, 2014). Furthermore, it has been shown that AD patients can suffer from confabulations (Attali et al., 2009; Cooper et al., 2006; Dalla Barba et al., 1999; El Haj and Larøi, 2017; Lee et al., 2007), and especially ‘habits’ confabulations (i.e. general habits incorrectly remembered as unique episodic event, La Corte et al., 2010). It has been shown that confabulation can be observed early in the AD continuum, notably resulting from a disconnection between frontal and temporal regions (Venneri et al., 2016). Confabulatory tendencies appear in early AD as a result of disconnection between crucial computational hubs in frontal and mediotemporal regions. By opposing AD participants with and without confabulations symptoms, we will be able to determine whether the severity of EM or executive symptoms can lead to the emergence of confabulations, or whether confabulations can be considered as an isolated symptom. Thus, we will try to determine which types of models seems to predict the best the cognitive deficits (executive, EM or a general cognitive failure) that leads to a confabulation syndrome.

## 2. Material and Methods

A total of 71 participants entered the study. They were all recruited at the Brain and Spine Institute in the Pitié-Salpêtrière University Hospital (Paris, France). Thirty-four fitted the clinical criteria for probable Alzheimer’s Disease (AD), ranging from mild to moderate AD, according to criteria of the Diagnostic and Statistical Manual of Mental Disorders (American Psychiatric Association, 2013) and the criteria of the National Institute of Neurological and Communicative Disorders and Stroke-Alzheimer’s Disease and Related Disorders Association (McKhann et al., 2011). Thirty-seven healthy control (HC) participants were recruited. HC were either spouses of patients or other individuals who volunteered to participate in the research projects of our laboratory. They had no history or current evidence of any medical, neurological, or psychiatric disease. They were also screened for any prior history of serious head injury, prolonged loss of consciousness, or active use of medications that could affect cognition or of recreational drugs. The study was conducted in accordance with the ethics standards laid down in the Declaration of Helsinki (2000) and was approved by the Ethical Committee of Île de France VI.

Demographical data are presented in the Table 1.

**Table 1.**
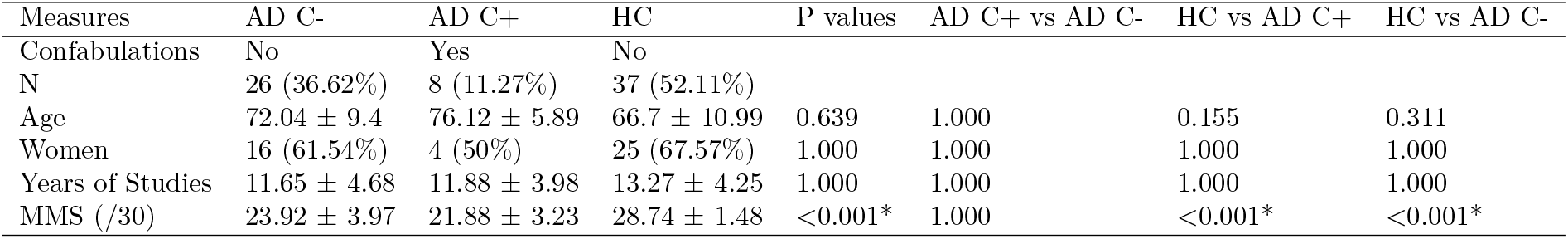
Demographic values of participants with and without confabulations. Notes : Mean ± Standard Deviation. AD = Alzheimer’s Disease. HC = Healthy Control. C+ = confabulatory syndrome. C-= Absence of a significant confabulatory syndrome.

| Measures | AD C- | AD C+ | HC | P values | AD C+ vs AD C- | HC vs AD C+ | HC vs AD C- |
| --- | --- | --- | --- | --- | --- | --- | --- |
| Confabulations | No | Yes | No |  |  |  |  |
| N | 26 (36.62%) | 8 (11.27%) | 37 (52.11%) |  |  |  |  |
| Age | 72.04 ± 9.4 | 76.12 ± 5.89 | 66.7 ± 10.99 | 0.639 | 1.000 | 0.155 | 0.311 |
| Women | 16 (61.54%) | 4 (50%) | 25 (67.57%) | 1.000 | 1.000 | 1.000 | 1.000 |
| Years of Studies | 11.65 ± 4.68 | 11.88 ± 3.98 | 13.27 ± 4.25 | 1.000 | 1.000 | 1.000 | 1.000 |
| MMS (/30) | 23.92 ± 3.97 | 21.88 ± 3.23 | 28.74 ± 1.48 | <0.001* | 1.000 | <0.001* | <0.001* |

### 2.1. Assessment of Confabulations

The presence of confabulations was assessed with Dalla Barba’s Confabulation Battery (CB) (Dalla Barba, 1993; Dalla Barba and Decaix, 2009; Dalla Barba et al., 2018a, 2018b). The CB is a questionnaire in which participants are asked to retrieve various information: episodic or semantic, past or to come, personal or general. It consists of 165 questions, 15 for each of the 11 following subfields: Personal Semantic Memory, Episodic Autobiographical Memory, Orientation in Time and Place, Linguistic Semantic Memory, Recent General Semantic Memory, Contemporary General Semantic Memory, Historical General Semantic Memory, Semantic Plans, Episodic Plans, ‘‘I don’t know” Semantic and Episodic questions. Regarding Episodic questions, participant’s answers are compared to his/her relatives’.

The 165 questions were presented in a semi-randomized order for balancing the 11 domains. Each answer was scored as ‘confabulation’, ‘wrong’, ‘I don’t know’ or ‘correct’ following the procedure already describe in previous works (La Corte et al., 2010).

Following the method that has been used in previous studies using the CB (Dalla Barba, 1993; Dalla Barba and Boisse, 2010; Dalla Barba and Decaix, 2009; Dalla Barba et al., 2018a; Serra et al., 2014), we considered the presence or absence of an ‘important confabulatory syndrome’ (C++) in participants who present at least 40% of “confabulation” responses on the EM (/Episodic Autobiographical Memory) questions. Likewise, participants who produce between 15% and 40% of confabulations would be considered as mild confabulators’ (C+). And finally, a participant with less than 20% of confabulation would be considered as non confabulators’ (C-). Thus, among our total population of 71 participants, no one can be consider as being C++, 8 met the C+ criteria, and all the remaining 63 were C-. All participants considered as ‘confabulators’ were AD subjects. Our sample thus consisted of 8 confabulators AD subjects (C+), 26 non-confabulators AD (C-), and 37 healthy control participants without confabulation.

### 2.2. Neuropsychological Examination

All participants underwent a comprehensive Neuropsychological Battery assessing global cognitive functioning, EM (in verbal and visual modalities), executive functioning, working memory and language. Global cognitive function was assessed with the Mini-Mental State Examination (MMSE) (Folstein et al., 1975).

EM assessment was assessed with two tests in order to distinguish between verbal and visual modalities. The verbal modality was assessed using the free and cued selective reminding test (FCSRT) (Grober and Buschke, 1987), a classical clinical memory test which has been showed to be particularly sensitive in detecting early memory decline (Di Stefano et al., 2015 ; Papp et al., 2017a), and specifically recommended for AD examination (Dubois et al., 2007, 2014; Epelbaum et al., 2017). It also offers the advantage to provide measurements of the main processes of EM (encoding, consolidation and retrieval) (Bertoux et al., 2013). Thus, using this test theoretically allows discriminating between a ‘pure’ memory deficit and a weakness in executives components of EM (e.g. strategic retrieval). The visual modality was assessed through the Rey-Osterrieth Complex Figure procedure (ROCF) (Fasteneau et al., 1999; Osterrieth, 1944) using a 3 minutes delay incident retrieval.

The ROCF immediate copy score was also used to assess instrumental abilities (e.g. visuo-spatial and praxis skills), as well as the Mill-Hill and the *Denomination* (Naming) 80 items (DO80; Deloche et al., 1989) (languages skills). The rest of the examination included the Raven’s PM47 (visuo-spatial reasoning); a forward and backward digit span (short term and working memory). Finally, executive functioning was tested with the Frontal Assessment Battery (FAB) (Dubois et al., 2000) and both phonemic and categorical Verbal Fluencies (Godefroy and GREFEX, 2008).

### 2.3. Statistical Analyses

Statistical analyses were perforxmed using R version 3.5.0 ([www.r-project.org]). The cognitive profiles of HC and AD patients were compared. Given the absence of significant group differences on demographic variables (see Table 1) on one hand, and the number of participants in our groups on the other hand, the Kruskall-Wallis non-parametric test was used at 5% significance level (p<0.05). Secondly, we used Pearson Partial correlation between the amount of confabulations (both in the EM field and the total number in the whole CB) and the neuropsychological performances. All *P-values* were corrected for multiple comparisons using the Benjamin-Hochberg method.

## 3. Results

There were no significant differences between our groups regarding the demographic variables (Table 1). The types of confabulations produced in the CB were compared between groups (see Table 2). Having been selected on this very criterion, confabulatory AD patients made significantly more confabulations for EM questions than non-confabulatory AD patients or HC participants (both p-values < 0.005). Furthermore, these patients also gave significantly more confabulatory answers than other groups for ‘Episodic Plans’ questions (both p-values < 0.005). These two domains are the only ones where confabulatory patients produced more confabulations than non-confabulatory AD patients. These patients also produced more confabulations answers than HC in ‘Personal Semantic’, ‘Recent General Semantic’ and overall in the whole battery (i.e. Total Confabulations proportion, all p values < 0.02). It has to be noted that the non-confabulatory AD group also produced significantly more confabulatory answers than the HC participants in the ‘Recent General Semantic’, ‘Episodic Memory’ and ‘Episodic Plans’ fields (all p values < 0.005). The distribution of the proportion of confabulatory answers in the 11 CB’s fields is presented in the Figure 1.

**Table 2.** Participants’ performance on the CB. Notes : Median (First Quartile [Q1], Third Quartile [Q3]). AD = Alzheimer’s Disease. HC = Healthy Control. C+ = confabulatory syndrome. C-= Absence of a significant confabulatory syndrome.

| Type | AD C+ | AD C- | HC | P values | AD C+ vs AD C- | HC vs AD C+ | HC vs AD C- |
| --- | --- | --- | --- | --- | --- | --- | --- |
| Personal Semantic Memory | 0 (0, 7.35) | 0 (0, 0) | 0 (0, 0) | 0.005* | 0.210 | 0.005* | 0.280 |
| Orientation in Time and Place | 0 (0, 3.33) | 0 (0, 0) | 0 (0, 0) | 0.806 | 1.000 | 0.806 | 0.806 |
| General Semantic Memory - Recent | 3.35 (0, 6.7) | 0 (0, 6.7) | 0 (0, 0) | 0.004* | 1.000 | 0.004* | 0.004* |
| General Semantic Memory - Historical | 0 (0, 0) | 6.7 (0, 11.65) | 0 (0, 6.7) | 0.280 | 0.410 | 1.000 | 0.292 |
| General Semantic Memory - Contemporary | 0 (0, 6.7) | 0 (0, 6.7) | 0 (0, 0) | 0.137 | 1.000 | 0.561 | 0.137 |
| Semantic Plans | 0 (0, 0) | 0 (0, 0) | 0 (0, 0) | 1.000 | 1.000 | 1.000 | 1.000 |
| Episodic Plans | 13.3 (11.65, 21.68) | 0 (0, 5.03) | 0 (0, 0) | <0.001* | 0.003* | <0.001* | 0.003* |
| Episodic Memory | 26.7 (20, 26.7) | 0 (0, 6.7) | 0 (0, 0) | <0.001* | <0.001* | <0.001* | 0.002* |
| Linguistic Semantic Memory | 0 (0, 0) | 0 (0, 0) | 0 (0, 0) | 0.627 | 1.000 | 0.627 | 0.627 |
| I don't Know Episodic | 0 (0, 0) | 6.7 (0, 13.3) | 6.7 (0, 6.7) | 0.365 | 0.365 | 0.365 | 1.000 |
| I don't Know Semantic | 6.7 (0, 16.65) | 6.7 (0, 13.3) | 13.3 (6.7, 13.3) | 1.000 | 1.000 | 1.000 | 1.000 |
| Total | 6.2 (5.17, 6.78) | 3.5 (2.4, 4.7) | 1.2 (1.2, 2.9) | 0.007* | 0.084 | 0.015* | 0.084 |

**Figure 1.**
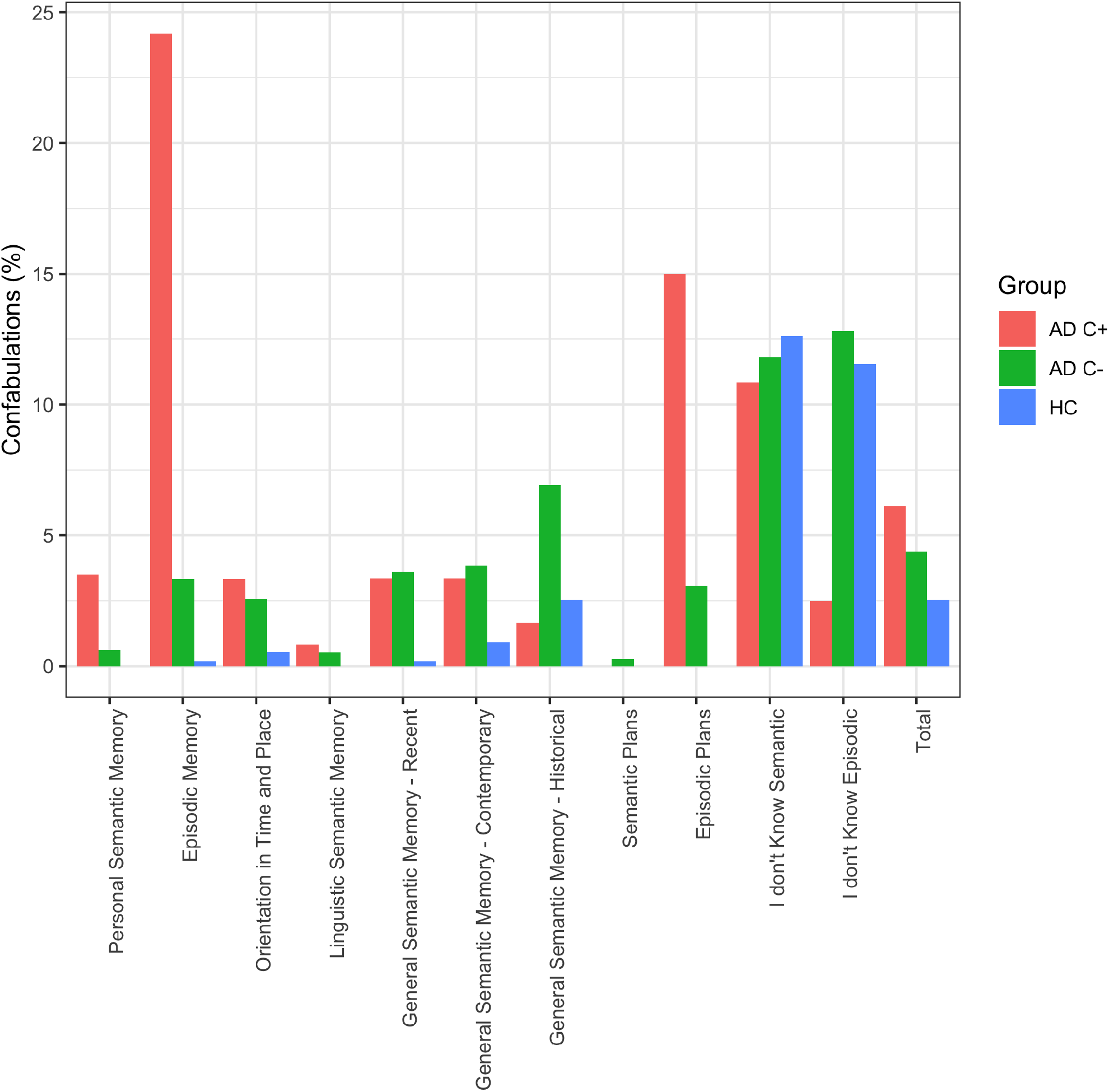
Proportion of Confabulations in the CB Fields by Groups. AD = Alzheimer’s Disease. HC = Healthy Control. C+ = confabulatory syndrome. C-= Absence of a significant confabulatory syndrome. *Tables*

Interestingly, regarding the neuropsychological performances (Table 3), there was no significant differences between our AD participants with and without confabulations (all p > 0.2). However, statistical analyses revealed significant differences between AD groups and HC participants on several cognitive tests, with HC participants systematically scoring better than patients. First, the general cognitive functioning, assessed via the MMSE score, was significantly better for our HC group than AD (*P values* < 0.001). The same can be observed on EM (i.e. FCSRT’s Free Recall – Immediate and Delayed –, Total Recall – idem –, Recognition score, ROCF Recall), executive (FAB) and language scores (Categorical Fluency and Naming). It has to be noted that significant differences were found between HC participants and non-confabulatory AD patients (but not with confabulatory AD patients) on some scores (i.e. PM47 and P Phonological Fluency), including the number of intrusions in FCSRT (this lack of difference with confabulators being mainly due to the significant variation in performance on this measure in this group).

**Table 3.** Cognitive Profiles of Confabulatory vs Non Confabulatory AD Participants. Notes : Median (Q1, Q3). AD = Alzheimer’s Disease. HC = Healthy Control. C+ = confabulatory syndrome. C-= Absence of a significant confabulatory syndrome, FCSRT = Free and Cued Reminding Selective Test, FAB = Frontal Assessment Battery, PM47 = Raven’s Colored Progressive Matrices.

| Measures | AD C+ | AD C- | HC | P values | AD C- vs AD C+ | HC vs AD C+ | HC vs AD C- |
| --- | --- | --- | --- | --- | --- | --- | --- |
| MMSE | 22.5 (21, 23.25) | 23.5 (21, 27.75) | 29 (28, 30) | <0.001* | 1.000 | <0.001* | <0.001* |
| Digit Span - Forward | 5 (5, 5) | 6 (5, 7) | 6 (5, 7) | 0.513 | 1.000 | 0.513 | 1.000 |
| Digit Span - Backward | 4 (3.5, 4.5) | 4 (3, 4) | 4 (4, 5) | 1.000 | 1.000 | 1.000 | 1.000 |
| FCSRT - Free Recall | 15 (10.5, 16.5) | 14 (9, 22) | 32.5 (27.25, 36) | <0.001* | 1.000 | 0.003* | <0.001* |
| FCSRT - Total Recall | 27 (20.5, 39.5) | 35 (28, 42) | 46.5 (45, 47) | <0.001* | 1.000 | 0.040* | <0.001* |
| FCSRT - Intrusions | 2 (2, 8) | 2 (1, 4) | 0 (0, 1) | 0.002* | 1.000 | 0.080 | 0.002* |
| FCSRT - Delayed Free Recall | 2 (0, 4) | 5 (1, 7) | 12.5 (11, 14) | <0.001* | 0.894 | 0.002* | <0.001* |
| FCSRT - Delayed Total Recall | 9 (4, 14.75) | 12 (10, 14) | 16 (16, 16) | <0.001* | 1.000 | <0.001* | <0.001* |
| Rey Figure - Memory Score | 3 (1, 10.5) | 7 (4.5, 14) | 17 (14.75, 24) | 0.002* | 1.000 | 0.010* | 0.010* |
| Rey Figure - Copy Score | 33 (30.75, 34) | 34 (29.75, 35) | 35 (33, 36) | 1.000 | 1.000 | 1.000 | 1.000 |
| FAB | 13 (11.5, 15) | 15 (13, 16) | 17 (16, 18) | 0.005* | 1.000 | 0.016* | 0.016* |
| Mill-Hill | 23.5 (23, 27) | 22.5 (14, 28.5) | 29 (25, 31) | 0.210 | 1.000 | 0.571 | 0.210 |
| PM47 Total | 23.5 (21.25, 25.75) | 22 (17.5, 25.5) | 32 (28, 34) | <0.001* | 1.000 | 0.054 | <0.001* |
| Categorical Fluency - Fruits | 7 (6.25, 10) | 11 (8.5, 12) | 16 (13.25, 18) | <0.001* | 0.229 | 0.003* | <0.001* |
| Categorical Fluency - Colors | 9 (7.5, 9.75) | 10 (9, 11) | 13.5 (11.25, 15) | <0.001* | 0.968 | 0.007* | <0.001* |
| Categorical Fluency - Birds | 6 (3.75, 6) | 6 (5, 10) | 13 (10, 15) | <0.001* | 1.000 | 0.002* | <0.001* |
| Litteral Fluency - P | 12 (7, 16) | 11 (8, 12) | 14.5 (13, 17) | 0.012* | 1.000 | 1.000 | 0.012* |
| Litteral Fluency - F | 7 (6, 9) | 8 (7, 12) | 12 (7.5, 15.75) | 1.000 | 1.000 | 1.000 | 1.000 |
| Litteral Fluency - L | 8 (7, 11) | 8 (7, 11) | 11 (9.25, 15) | 0.612 | 1.000 | 1.000 | 0.612 |
| Naming (BECS - GRECO) | 37 (37, 38) | 38 (31.5, 38.75) | 40 (39, 40) | 0.001* | 1.000 | 0.010* | 0.003* |

**Table 4.** Correlation Between Confabulations and Cognitive Scores. Notes : FCSRT = Free and Cued Reminding Selective Test, FAB = Frontal Assessment Battery, PM47 = Raven’s Colored Progressive Matrices.

| Measures | Total Confabulations | EM Confabulations |
| --- | --- | --- |
| MMSE | -0.83 | -0.55 |
| Digit Span - Forward | -0.91 | -0.69 |
| Digit Span - Backward | -0.87 | -0.62 |
| FCSRT - Free Recall | -0.89 | -0.6 |
| FCSRT - Total Recall | -0.93 | -0.74 |
| FCSRT - Intrusions | -0.89 | -0.68 |
| FCSRT - Delayed Free Recall | -0.65 | -0.57 |
| FCSRT - Delayed Total Recall | 0.75 | 0.62 |
| Rey Figure - Memory Score | -0.73 | -0.53 |
| Rey Figure - Copy Score | 0.87 | 0.69 |
| FAB | 0.86 | 0.68 |
| Mill-Hill | 0.89 | 0.66 |
| PM47 Total | -0.47 | -0.55 |
| Categorical Fluency - Fruits | 0.88 | 0.63 |
| Categorical Fluency - Colors | -0.73 | -0.5 |
| Categorical Fluency - Birds | -0.92 | -0.7 |
| Litteral Fluency - P | 0.53 | 0.28 |
| Litteral Fluency - F | 0.91 | 0.7 |
| Litteral Fluency - L | 0.39 | 0.31 |
| Naming (BECS - GRECO) | -0.54 | -0.3 |

Finally, there were no significant correlations between the number of confabulations (both Episodic and in the entirety of the CB) and the neuropsychological scores of different cognitive functions.

## 4. Discussion

The aim of our study was to determine the cognitive profile of confabulatory patients, using AD participants with and without confabulations opposed on a comprehensive neuropsychological battery. The present results demonstrate an absence of difference in the cognitive profile of AD participants with and without a confabulatory syndrome, both of them demonstrating lower cognitive performances than the HC group in the EM and executive domains as it could be expected from literature (Dubois et al., 2007; Rajan et al., 2015; Schmid et al., 2013). This statistically equivalent cognitive profile between our two groups questioned some models of confabulation, which assume a unique and sufficient cognitive (e.g. executive) process underlying the onset of confabulations.

In the confabulation literature, at least three type of models have been identified depending of which type of cognitive processes is thought to underlie the appearance of this symptom (Glowinski et al., 2008): executive failure, temporal ordering deficit, and multi-factorials models. In our work, participants were assessed with both executive and episodic tests. Our objective was, by comparing the cognitive profile of participants with and without confabulation, to determine whether or not there would be a difference in one or both of these cognitive domains. Thus, an executive deficit in confabulatory, but not in non confabulatory participants would support the first presented approach, while a significant difference in EM performances would support the second one. HC and mild to moderate AD participants entered statistical analyses. Since the cognitive profile of typical AD is well known (Dubois et al., 2007, 2014), a variation from that profile would be easily detected.

The first approach proposes a retrieval dysfunction, with two different cognitive mechanisms involved in confabulation (Metcalf et al., 2007): one centered on a source monitoring process (Johnson and Raye, 1998; Johnson et al., 1993), and another focusing on a deficient strategic retrieval process (Moscovitch and Melo, 1997). Both these retrieval approaches suppose an executive origin of confabulations. Executive functions are an ensemble of high-level cognitive processes allowing goal-directed tasks or behavior regulation and are highly – although not exclusively – related to the frontal cortex integrity (Godefroy et al., 2010; Seniów, 2012). In memory, executive functions are involved in both encoding and retrieval (Putcha et al., 2018), involving PFC activity (Cabeza, 2002; Habib et al., 2003). Regarding, confabulations, the executive approach of confabulations is supported by two types of arguments at neuronanatomical and cognitive level. On the neuronanatomical side, it has been shown that the PFC seems to play a crucial role in the occurrence of confabulations (Moscovitch and Melo, 1997). For example, reviewing 79 cases of patients experiencing confabulations with various etiologies, Gilboa and Moscovitch (2002) showed that 81% (i.e. 64) had a PFC lesion (either right or left). On a cognitive aspect, some previous results have shown some co-occurrences and correlations between executive troubles and confabulations. For example, the amount of perseverations (Kopelman et al., 1997). In their model of confabulation, Gilboa and Moscovitch proposed that the PFC, and more specifically the ventromedial area, would mediate confabulations by supporting the ability to generate context-relevant scheme and to compare it to the stocked memories (Gilboa and Moscovitch, 2002, 2017; Moscovitch and Melo, 1997). Source monitoring processes have not been specifically assessed in our study. Yet, regarding strategic retrieval, we used some EM and executive tests involving this cognitive process. Indeed, the FCSRT is often used in neuropsychological assessment because allowing distinguishing between the three memory phases: encoding, consolidation and retrieval. In detail, a retrieval deficit would correspond to a weakness of FR with a normalization of performance in TR through semantic cueing. Moreover, for executive functioning, the fluency task’s performances (especially the phonological condition) are driven by executive strategic processes (Luo et al., 2010; Shao et al., 2014). Given that these scores are not found statistically different between our groups, nor correlated with the confabulatory answers proportion, in our sample confabulation can not be traced back to a strategic retrieval deficit. These results are coherent with previous observations of patients with confabulations, without executive deficits (Dalla Barba, 1993; Dalla Barba et al., 1999; Nedjam et al., 2004).

Another theoretical approach rather proposes that confabulations could result from multiple cognitive deficits (Glowinski et al., 2008). Those theoretical frameworks generally propose that confabulations, and especially spontaneous confabulations, would rise from the convergence of EM and executive deficits For example, Metcalfe and colleagues (2007) proposed that executive deficits (i.e. retrieval and evaluation deficits) could influence the emergence of confabulations, and its content could be determined by memory and emotional/motivational personal biases. Regarding the general cognitive state of our groups, the AD participants did not statistically differ in the global cognitive performance whether they present confabulations or not (i.e. MMSE). Regarding the convergence of both executive and EM troubles, there were no differences between our two groups on any of these measures.

Finally, the last theoretical approach postulates a temporal ordering dysfunction underlying confabulations (Dalla Barba, 1993, 2002; Dalla Barba and Boisse, 2010; Dalla Barba and La Corte, 2013), assuming that an executive / frontal deficit would not be necessary. Within this framework, the authors propose to see confabulations as misplacement of real memory elements in an inadequate context. According to the Memory, Consciousness and Temporality Theory (MCTT, Dalla Barba, 2002), confabulatory patients would mistake a general memory scheme as a specific event (Attali et al., 2009; Dalla Barba, 2002; Dalla Barba and Boisse, 2010) as a consequence of a Temporal Consciousness (TC) dysfunction (La Corte et al., 2010). Usually, TC allows individuals to be oriented in the present, remember the personal past and project the personal future. While confabulatory patients still have a TC — otherwise they could not recall any event doing mental time travel, even a wrong one —, it is interacting with stables patterns of modifications of the nervous systems instead of those tied to specific contextual events (Dalla Barba and Boisse, 2010; Dalla Barba and La Corte, 2015; La Corte et al., 2011a, 2011b). Dalla Barba and La Corte propose that the hippocampus is the anatomical correlate of TC (Dalla Barba and Kopelman, 2017; Dalla Barba and La Corte, 2013). One prediction of this approach is that confabulatory patients would not only be affected in their capacity to recall their personal memories from the past (i.e. EM), but would also produce erroneous statements about their present situation and even when asked to predict their future (Dalla Barba, 2002). In our study, confabulatory participants produce significantly more confabulations not only in the EM domain (which was the group inclusion criteria), but also for ‘Episodic Plans’ questions, showing that confabulation often involves personal temporality as a whole (see Figure 1). These results are in line with the MCTT predictions. Our sample was composed of mild to moderate AD patients, a pathology which produce a progressive hippocampal atrophy (Dubois et al., 2007, 2014; Scheltens et al., 2016). We could thus expect that the evolution of the hippocampal atrophy would not only make EM performances fall, but also prevent patients to confabulates, evolving from disruption to abolition of TC. Regarding the EM scores, we used two classical neuropsychological tests, i.e. the FCSRT and the ROCF, respectively assessing the verbal and visual modality of EM. In the same way as for executive functioning, none of the EM neuropsychological scores differed between our two AD groups, suggesting no implication or link between the EM processses and the tendency to produce provoked confabulations.

Taken together, our result suggests that, in our population, the occurrence of a confabulatory syndrome does not depend of the presence or absence of a single dysexecutive syndrome or EM deficit. This goes both against the models, which assume a sufficient origin of executive, or EM troubles in the occurrence of confabulation.

Despite the absence of distinction of our AD participants with or without confabulations, group differences were found in our data indicating, consequently and as expected, an opposition between AD and HC participants.

First of all, no group effect for instrumental functions (except for Naming, a difference that could reflect a lexical-semantic weakness known to be present in AD, see below). This is relevant with the pathological stage (i.e. mild to moderate) of our AD groups (Wang et al., 2018; Yesavage et al., 1993).

A significant group effect was found for some EM and executive measures. For executive measures, this difference concerned categorical, but no literal, fluency tasks and the PM47 total score. Regarding the fluency task, despite both literal and categorical fluency include a frontal / executive load, the two can be distinguish (Costafreda et al., 2006; Vonk et al., 2019) with the categorical fluency cares and additional semantic component (Shao et al., 2014). This result is coherent with previous literature demonstrating an alteration of the semantic domain, even at early stages of the course of AD (Amieva et al., 2008; Gonzalez-Nosti et al., 2018; Grossman et al., 2003; Papp et al., 2017b).

For EM measures, a significant group effect and differences between AD and HC participants were found in all measures except for immediate recall. First, the absence of difference in Immediate Recall is not surprising as AD mostly affect EM, which has to be distinguished from immediate recall as it recruit different processes and brain network (Casaletto et al., 2017). Regarding EM differences, again those results fit with literature and what is known about the cognitive profile of AD (Dubois et al., 2007, 2014; Sarazin et al., 2007). Indeed, it has been shown that, over the course of AD, the FR measure would start to decline earlier than for TR (Grober et al., 2000, 2018), which would be a marker of presence of amyloidosis (Papp et al., 2017a; Schindler et al., 2017), expressing an encoding/retrieval memory processes (Papp et al., 2017a). Possibly, differences were present in our data concerning intrusions. Like confabulations, intrusions belong to what some authors call ‘positive’ EM disorders, i.e. memory distortions and incorrect productions (Attali et al., 2009; Dalla Barba and Wong, 1995; Dalla Barba et al., 1995). In the FCSRT, intrusions are defined as the production, in FR or TR, of a non-studied item. These difficulties are part of the classical neuropsychological and EM profile of AD (Dubois et al., 2007, 2014), present very early in the disease (Loewenstein et al., 2018; Thomas et al., 2018), and have been shown to be highly discriminant between AD and other neurodegenerative diseases (Teichmann et al., 2017). As our participants are at a mild to moderate stage of AD, this difference again fit well with previous literature.

To summarize, concerning the differences between our AD and HC participants, our results are consistent with previous knowledge about AD cognitive profile.

Altogether, HC participants produced few confabulations, but there was two domains in which they tended to produce more confabulatory answers, namely the “I don’t know” questions. These results also confirm previous research using the CB in healthy people (Dalla Barba et al., 2018b). Dalla Barba and colleagues (2018b) explained that this tendency could be explained by a natural inclination to answer the questions, even when the answer is not available.

Nevertheless, this study have some limitations, notably the confabulatory patients sample with only 8 AD patients presenting a confabulatory syndrome. Moreover, our group only includes patients with a moderate confabulatory syndrome. Indeed, confabulations are not a very frequent symptom in AD patients. Thus, as Venneri, Mittolo and Marco (2016) observed, this requires to be careful in the generalization of these conclusions regarding confabulations in general.

To conclude, in our study, in agreement with different models of confabulations, we investigated whether a confabulatory syndrome in mild to moderate AD could emerge from a unique EM or executive syndrome. In demonstrating an absence of statistical difference between our participant’s cognitive profile depending on the presence or absence of confabulations, our findings seem to put in doubt the cognitive models of confabulations that postulates that these symptoms would emerge from an isolated executive disorder. On the other hand, demonstrating that confabulatory participants significantly produce more Episodic confabulations, regarding both past and future dimensions, our results support a temporal consciousness account of confabulation.

## Data Availability

All data produced in the present study are available upon reasonable request to the authors

